# Oral Premedication with Tapentadol versus Pregabalin for Acute Postoperative Pain in Lower Limb Surgery Under Neuraxial Anesthesia: A Randomised Controlled Trial

**DOI:** 10.64898/2026.05.14.26353251

**Authors:** Ma. del Rosario Escalona-Arroyo, Phabel A. Lopez-Delgado, Mirna M. Delgado-Carlo

**Author notes:** **Corresponding author:** Mirna M. Delgado-Carlo MD, **Email:****, **. **First Author:** Ma. del Rosario Escalona-Arroyo MD, **Email:**. **Co-Author:** Phabel A. Lopez-Delgado BSc, **Email:**.

## Abstract

**Background:** Acute postoperative pain affects more than 80% of surgical patients, with orthopedic lower limb procedures consistently associated with severe pain intensity and high opioid requirements. Preemptive analgesia with oral agents has been proposed to attenuate central and peripheral sensitization prior to surgical incision. Tapentadol, a dual-mechanism *µ*-opioid receptor agonist and norepinephrine reuptake inhibitor, and pregabalin, a voltage-gated calcium channel modulator, represent pharmacologically distinct premedication options; however, direct comparative data in this surgical context are lacking. This randomized controlled trial compared the analgesic efficacy and safety of 72-hour oral premedication with tapentadol versus pregabalin in patients undergoing elective lower limb surgery under neuraxial anesthesia.

**Methods:** In this double-blind, parallel-group trial, 46 patients undergoing elective lower limb surgery under neuraxial anaesthesia were randomized to tapentadol 50 mg q12h or pregabalin 75 mg q24h for 72 h preoperatively. The sample size was calculated using a two-sided test for comparing two proportions, with *α* = 0.05 and 80% power, assuming a 30% difference in the proportion of patients requiring rescue analgesia between groups (70% vs 40%), yielding 23 patients per group; accounting for 10% attrition, 46 patients were enrolled. Pain intensity was assessed using the Numeric Rating Scale (NRS, 0–10) and Verbal Rating Scale (VRS) at PACU arrival (T0) and at 30 (T1), 60 (T2), 90 (T3), and 120 (T4) minutes thereafter. Secondary outcomes included rescue morphine consumption and safety. Between-group comparisons used a linear mixed model.

**Results:** Pain scores diverged from 30 minutes onward, with a large observed difference at 90 minutes favouring tapentadol (mean NRS difference –0.91, p = 0.006, Cohen’s d = –0.96). Rescue morphine was required in 4.3% of tapentadol patients versus 21.7% of pregabalin patients. Nausea and vomiting occurred in 17.4% of both groups; no hypersensitivity reactions were observed.

**Conclusions:** Seventy-two-hour oral premedication with tapentadol 100 mg/day provided superior postoperative analgesia compared with pregabalin 75 mg/day at the 90-minute PACU timepoint, with a large effect size and a fivefold reduction in rescue analgesia requirements. Both agents were well tolerated. These findings support the use of tapentadol as a premedication strategy in lower limb orthopaedic surgery.

**Trial registration:** https://ClinicalTrials.gov - NCT07587645 (retrospectively registered).

**Highlights:**

1. RCT (n=46) comparing 72-hour oral premedication with tapentadol versus pregabalin in lower limb surgery.
2. Tapentadol provided superior analgesia at 90 minutes (Cohen’s d = 0.96, large effect).
3. Rescue analgesia was required in 4.3% (tapentadol) vs. 21.7% (pregabalin).
4. No hypersensitivity reactions; both agents were well tolerated.
5. These findings support the use of tapentadol as a premedication strategy in lower limb orthopaedic surgery.

**Figure 1: Graphical Abstract – CONSORT:** Figure 1:
CONSORT flow diagram.
Enrollment, allocation, and analysis of participants in the pilot randomized controlled trial comparing oral premedication with tapentadol versus pregabalin for acute postoperative pain in elective lower limb surgery under neuraxial anesthesia.

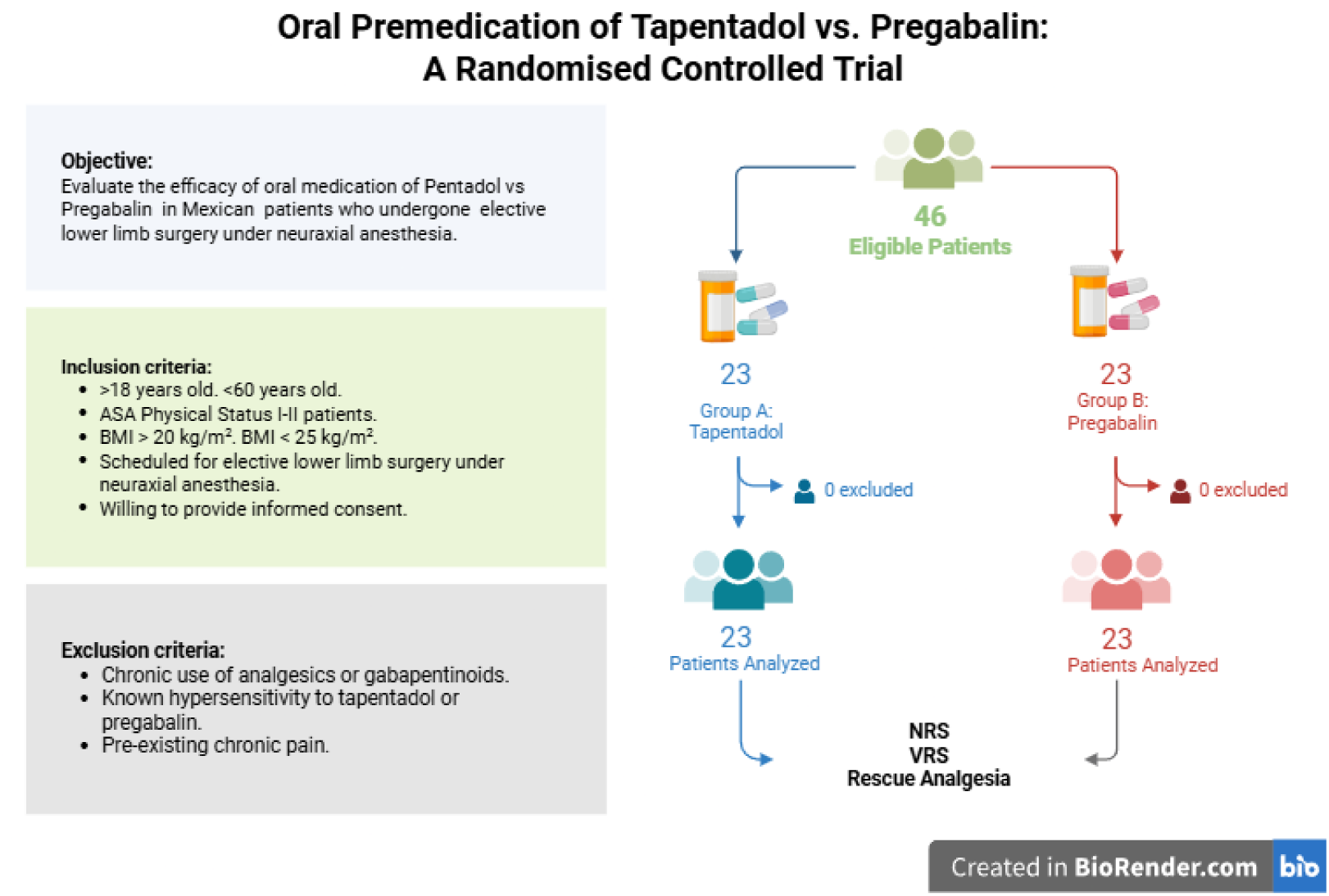

## 2. Introduction

Acute postoperative pain (APP) remains one of the most prevalent and undertreated complications of surgical care, affecting more than 80% of patients undergoing surgical procedures, of whom approximately 75% rate their pain as moderate to severe [1, 2]. Beyond patient suffering, inadequately controlled APP drives a cascade of adverse physiological responses, including sympathetic activation, impaired respiratory function, and neuroendocrine stress, that increase perioperative morbidity, prolong hospital stay, and raise the risk of transition to chronic postoperative pain [3, 4]. Orthopaedic procedures involving the lower limb are particularly associated with severe APP, as bone and periosteum carry the lowest nociceptive threshold among deep somatic structures, and patients undergoing such surgery consistently report higher pain scores and greater opioid requirements than those undergoing abdominal or soft-tissue procedures [5].

Preemptive analgesia, the administration of analgesic agents prior to surgical incision, has been proposed as a strategy to attenuate central and peripheral sensitisation before the nociceptive barrage of surgery is established, thereby reducing APP intensity and opioid consumption in the postoperative period [4, 6]. Two agents with distinct but complementary mechanisms of action are of particular interest in this context. Pregabalin, a structural analogue of *γ*-aminobutyric acid (GABA), binds to the *α*_2_-*δ* subunit of voltage-gated calcium channels, dampening neuronal excitability and attenuating central sensitisation; its perioperative use has been associated with reductions in postoperative pain scores and opioid consumption [6, 7]. Tapentadol is a centrally acting analgesic with a dual mechanism: agonism at the *µ*-opioid receptor and inhibition of norepinephrine reuptake, two actions that interact synergistically to produce analgesia across both nociceptive and neuropathic pain states [8, 9]. Unlike conventional opioids, tapentadol does not rely on active metabolites and exhibits a more predictable pharmacokinetic profile with a comparatively favourable gastrointestinal tolerability [8]. Despite growing clinical interest in both agents, direct comparative data on their efficacy as oral premedication for APP in lower limb orthopaedic surgery under neuraxial anesthesia remain scarce.

We conducted a randomized controlled trial to compare the analgesic efficacy of oral premedication with tapentadol (50 mg every 12 hours for 72 hours preoperatively) versus pregabalin (75 mg every 24 hours for 72 hours preoperatively) in patients undergoing elective lower limb surgery under neuraxial anesthesia. The sample size was calculated using a two-sided test for comparing two proportions, with *α* = 0.05 and 80% power, assuming a 30% difference in the proportion of patients requiring rescue analgesia between groups (70% in the tapentadol group vs 40% in the pregabalin group). This yielded 23 patients per group; accounting for 10% attrition, a total of 46 patients were enrolled. Pain intensity was assessed with the Numeric Rating Scale (NRS) and the Verbal Rating Scale (VRS) at five timepoints during the first two hours of post-anesthesia care unit (PACU) recovery. We hypothesised that tapentadol would provide superior analgesia compared to pregabalin. The study was conducted at the Hospital Regional “General Ignacio Zaragoza,“ ISSSTE, Mexico City (institutional prospective registry RPI #386-2024) and retrospectively registered at https://ClinicalTrials.gov (NCT07587645).

## 3. Methods

### 3.1. Study design and ethics

This was a single-centre, prospective, randomized, controlled, parallel-group trial conducted at the Hospital Regional “General Ignacio Zaragoza,“ ISSSTE, Mexico City. The study was approved by the institutional Ethics and Research Committee and conducted in accordance with the Declaration of Helsinki and Mexican national research regulations (*Ley General de Salud*, Título Segundo) [10]. Written informed consent was obtained from all participants prior to enrolment.

### 3.2. Sample Size Calculation

The sample size was calculated using a two-sided test for comparing two proportions, with *α* = 0.05 and 80% power. Assuming a 30% difference in the proportion of patients requiring rescue analgesia between the tapentadol group (70%) and the pregabalin group (40%), the required sample size was 23 patients per group. Accounting for a 10% attrition rate, the target sample size was set at 46 patients (23 per group).

### 3.3. Participants

Eligible patients were aged 18–50 years, ASA physical status I or II, with a body mass index between 18 and 35 kg/m^2^, scheduled for elective lower limb surgery under neuraxial anesthesia. Key exclusion criteria were: chronic use of analgesics or gabapentinoids, known hypersensitivity to tapentadol or pregabalin, and pre-existing chronic pain. Elimination criteria included hypersensitivity reaction to any study drug during administration, hemodynamic shock of any etiology, requirement for advanced airway management or post-operative mechanical ventilation, surgical duration exceeding 120 minutes, requirement of an epidural catheter dose within the first two postoperative hours, and voluntary withdrawal of consent at any point.

### 3.4. Randomization and blinding

A total of 46 patients were allocated equally to two parallel groups (*n* = 23 for each one) using simple random allocation with sequentially numbered assignments. Group A (TAP) received tapentadol 50 mg orally every 12 hours, and Group B (PREG) received pregabalin 75 mg orally every 24 hours, both initiated 72 hours before the scheduled surgical incision. Study drugs were prepared and dispensed in identical oral formulations by personnel not otherwise involved in patient care or outcome assessment, ensuring blinding of both the patient and the treating anaesthesiologist. Neither the patients nor the investigator assessing pain outcomes had knowledge of group assignment throughout the study period.

### 3.5. Anesthetic Protocol

On arrival to the operating theatre, standard non-invasive monitoring was established (electrocardiography, pulse oximetry, non-invasive blood pressure). Supplemental oxygen was administered at 1.5 L/min via nasal cannula. Neuraxial anesthesia was performed with the patient in the lateral decubitus position, following aseptic technique. After skin infiltration with lidocaine 2% (60 mg), a Tuohy #17 needle was introduced at the L2–L3 intervertebral space using the loss-of-resistance technique; a Whitacre #27 spinal needle was then advanced through the Tuohy needle, and bupivacaine 150–200 *µ*g/kg was administered intrathecally upon confirmed cerebrospinal fluid return. Metameric spread to a minimum of T10 and maximum of T8 was verified before proceeding. During the intraoperative period, paracetamol 1 g IV was administered as part of the maintenance analgesic regimen; opioid analgesics were withheld, and no subsequent neuraxial doses were administered via epidural catheter.

### 3.6. Outcomes

The primary outcome was postoperative pain intensity, assessed using the Numeric Rating Scale (NRS, 0–10) and the Verbal Rating Scale (VRS: 0 = absence; 1 = low; 2 = moderate; 3 = severe) [11, 12] at five timepoints: post-anesthesia care unit (PACU) arrival (T0), and at 30 (T1), 60 (T2), 90 (T3), and 120 (T4) minutes thereafter.

Secondary outcomes included the number of rescue analgesia doses, total morphine consumption (mg), and incidence of nausea and vomiting. Rescue analgesia (morphine 4 mg IV) was administered as a single dose on patient request or clinical indication. Hypersensitivity reactions were classified according to Müller criteria and managed per institutional protocol (hydrocortisone 100 mg IV; adrenaline 0.5 mg IM for anaphylaxis) [13].

### 3.7. Statistical analysis

Descriptive statistics are reported as mean *±* standard deviation (SD) for normally distributed continuous variables and as frequencies with percentages for categorical variables. Baseline between-group comparability was assessed using the Mann–Whitney *U* test for continuous variables and Pearson’s chi-squared test for categorical variables.

The primary analysis modelled the longitudinal NRS trajectory using a linear mixed model (LMM) with Group, Time (T0–T4 as a categorical factor), and their interaction (Group *×* Time) as fixed effects, and a random intercept per patient to account for within-subject correlation across repeated measurements. The model was fitted by restricted maximum likelihood (REML) using the lme4 package, with Satterthwaite-approximated degrees of freedom and *p*-values obtained via lmerTest [14]. Post-hoc between-group contrasts at each timepoint were derived using estimated marginal means (emmeans package) with Holm correction for multiple comparisons; effect sizes are reported as Cohen’s *d* computed from the model residual standard deviation.

A sensitivity analysis repeated the primary model with time entered as a continuous variable (minutes: 0, 30, 60, 90, 120) to estimate between-group differences in the rate of pain increase. VRS and rescue analgesia data were analysed using Pearson’s chi-squared test.

All analyses were performed in R version 4.5.3, using the lme4 and lmerTest packages for the primary LMM [14] and the ggstatsplot package for exploratory between-group comparisons [15]. A two-sided *p <* 0.05 was considered statistically significant.

### 3.8. Trial registration

This trial was prospectively registered with the institutional research registry of the Instituto de Seguridad y Servicios Sociales de los Trabajadores del Estado (ISSSTE) under registration number **RPI #386-2024**. The trial was subsequently retrospectively registered on *https://ClinicalTrials.gov* (**NCT07587645**); as the study was conducted prior to the availability of an international registry to the investigative team, registration was completed after data collection and is reported transparently in accordance with CONSORT 2025 guidance on retrospective registration disclosure.

## 4. Results

### 4.1. Recruitment and Retention

A total of 46 patients were enrolled over a one-month recruitment period (9.2 patients per month). All 46 participants completed the 120-minute follow-up (retention 100%), and every patient received the allocated intervention without major protocol deviations (adherence 100%).

### 4.2. Participant Characteristics

Forty-six patients were enrolled and completed the study (23 in each group); no participants were eliminated after randomization, and no missing data were present. Baseline characteristics are presented in Table 1. Groups were comparable across all demographic and clinical variables. Mean age was 35 years in the pregabalin group, and 34.2 years in the tapentadol group. The tapentadol group had a mean weight of 74.3 kg and mean BMI of 26.1 kg/m^2^; the pregabalin group had a mean weight of 76.3 kg and mean BMI of 27.0 kg/m^2^. ASA physical status I was predominant in both groups (tapentadol: 69.6%; pregabalin: 65.2%). Mean surgical duration was 72 minutes (range 35–107 minutes) in the tapentadol group and 84 minutes (range 39–117 minutes) in the pregabalin group, with no statistically significant difference between groups (p = 0.08). The majority of procedures were performed by the traumatology and orthopaedics service (85%), with the remainder distributed across general surgery (11%), plastic surgery (3%), and surgical oncology (1%). All patients presented with NRS = 0 and VRS = “absence” at PACU arrival (T0), confirming homogeneous baseline pain status.

**Table 1:** Baseline characteristics by treatment group.

| Variable | Tapentadol<br>(100 mg/day; $n = 23$) | Pregabalin<br>(75 mg/day; $n = 23$) | $p$-value |
| --- | --- | --- | --- |
| Age, mean (range), yr | 34.2 (18–50) | 35.0 (18–50) | 0.77 |
| Sex, M/F | 14/9 | 15/8 | 0.76 |
| ASA I/II | 16/7 | 15/8 | 0.75 |
| Weight, mean (range), kg | 74.3 (56–91) | 76.3 (56–93) | 0.57 |
| BMI, mean (range), kg/m<sup>2</sup> | 26.1 (21.3–30.1) | 27.0 (23.2–31.5) | 0.30 |
| Surgical duration, mean (range), min | 72 (35–107) | 84 (39–117) | 0.08 |
| NRS at T0 | 0 (all) | 0 (all) | n/a |
| VRS at T0 | Absence (all) | Absence (all) | n/a |

ASA = American Society of Anesthesiologists physical status; BMI = body mass index; NRS = Numeric Rating Scale; VRS = Verbal Rating Scale. $p$ -values from Mann–Whitney $U$ test (continuous variables) and Pearson’s chi-squared test (categorical variables).

### 4.3. Primary Outcome: NRS Pain Trajectory

NRS pain scores over time are shown in Figure 2. At T0, both groups were pain-free (NRS = 0 in all patients). From T1 onward, pain scores diverged progressively, with the pregabalin group reporting consistently higher NRS values at every subsequent timepoint.

**Figure 2:**
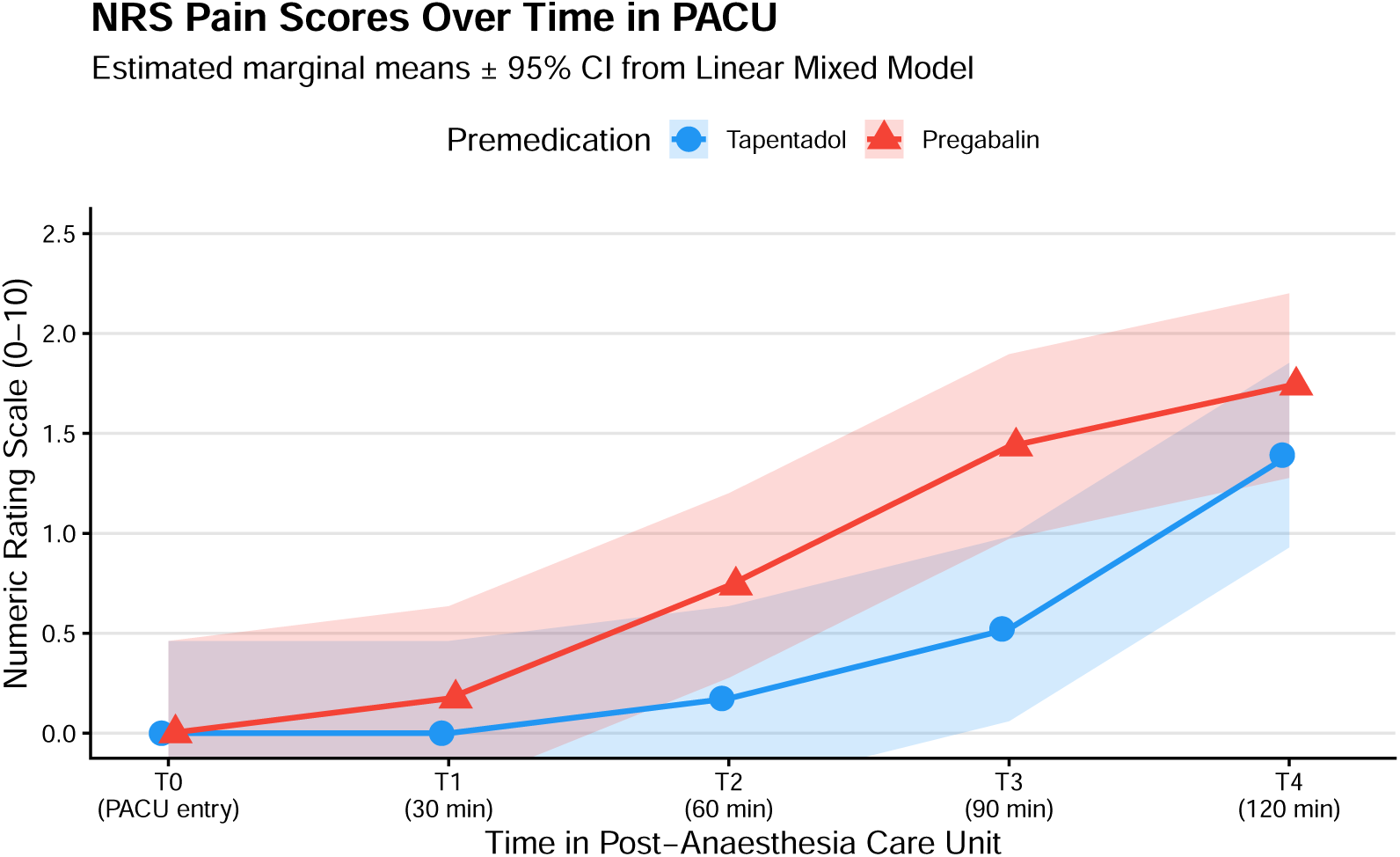
NRS pain trajectory over time by treatment group. Estimated marginal means *±* 95 % confidence intervals derived from the primary linear mixed model. Pain scores were identical at T0 (NRS = 0) in both groups and diverged progressively from 30 minutes onward. A statistically significant between-group difference was observed at T3 (90 min; mean difference *−*0.91, *p* = 0.006, Cohen’s *d* = 0.96). TAP = tapentadol; PREG = pregabalin; PACU = post-anesthesia care unit; NRS = Numeric Rating Scale.

The primary linear mixed model (LMM) revealed a highly significant main effect of Time (*F*_4,181.6_ = 23.61, *p <* 0.001), confirming that pain increased over the two-hour PACU observation window in both groups. The Group *×* Time interaction did not reach statistical significance at the overall model level (*F*_4,181.6_ = 1.75, *p* = 0.142), which is consistent with the sample size and the limited power of a 46-patient study to detect interaction effects. However, a sensitivity analysis modeling time as a continuous variable demonstrated a borderline-significant Group *×* Time interaction (*F*_1,187.6_ = 3.79, *p* = 0.053), indicating that the rate of pain increase over the PACU period was faster in the pregabalin group.

Post-hoc between-group contrasts at each timepoint, derived from estimated marginal means with Holm correction, are presented in Table 2. At T3 (90 minutes), tapentadol patients reported significantly lower pain scores than pregabalin patients (mean difference *−*0.91 NRS units, 95% CI [*−*1.56, *−*0.26]; *t*_168.1_ = *−*2.73, *p* = 0.006; Cohen’s *d* = *−*0.96, large effect). A medium-effect trend was observed at T2 (60 minutes; mean difference *−*0.56, *p* = 0.089, *d* = *−*0.59). At T4 (120 minutes), scores began to converge as neuraxial block regression neared completion and ward analgesia was initiated (mean difference *−*0.34, *p* = 0.294, *d* = *−*0.37).

**Table 2:** Between-group NRS contrasts at each PACU timepoint (Tapentadol vs. Pregabalin)

| Timepoint | Mean<br>NRS<br>diff. | 95 % CI | SE | $t$ (tau) | $p$ (Holm) | Cohen’s<br>$d$ |
| --- | --- | --- | --- | --- | --- | --- |
| T0 (0 min) | 0.00 | (−0.65, 0.65) | 0.33 | 0.00 | 1.000 | 0.00 |
| T1 (30 min) | −0.17 | (−0.82, 0.47) | 0.33 | −0.53 | 0.599 | −0.18 |
| T2 (60 min) | −0.56 | (−1.21, 0.08) | 0.33 | −1.71 | 0.089 | −0.59 |
| T3 (90 min) | −0.91 | (−1.56, −0.26) | 0.33 | −2.76 | 0.006 | −0.96 |
| T4 (120 min) | −0.34 | (−1.00, 0.30) | 0.33 | −1.05 | 0.294 | −0.37 |

Estimates from a linear mixed model with Group, Time, and Group × Time interaction as fixed effects and random intercept per patient. Contrasts computed via `emmeans` with Holm correction. Cohen’s $d$ is calculated from the model residual SD. Negative values favour tapentadol. PACU = post-anesthesia care unit; SE = standard error; TAP = tapentadol; PREG = pregabalin.

### 4.4. Secondary Outcome: Verbal Rating Scale

VRS findings were consistent with the NRS data (Figure 3). All patients in both groups rated pain as “absence” at T0. The proportion of patients reporting any pain increased progressively from T1 onward, and was higher in the pregabalin group at each timepoint. At T3, in the tapentadol group, 73.9% reported no pain and 26.1% reported mild pain; no tapentadol patient reported moderate pain. In the pregabalin group, 56.5% reported no pain, 30.4% mild pain, and 13.0% moderate pain. No patient in either group reported severe pain at any timepoint.

**Figure 3:**
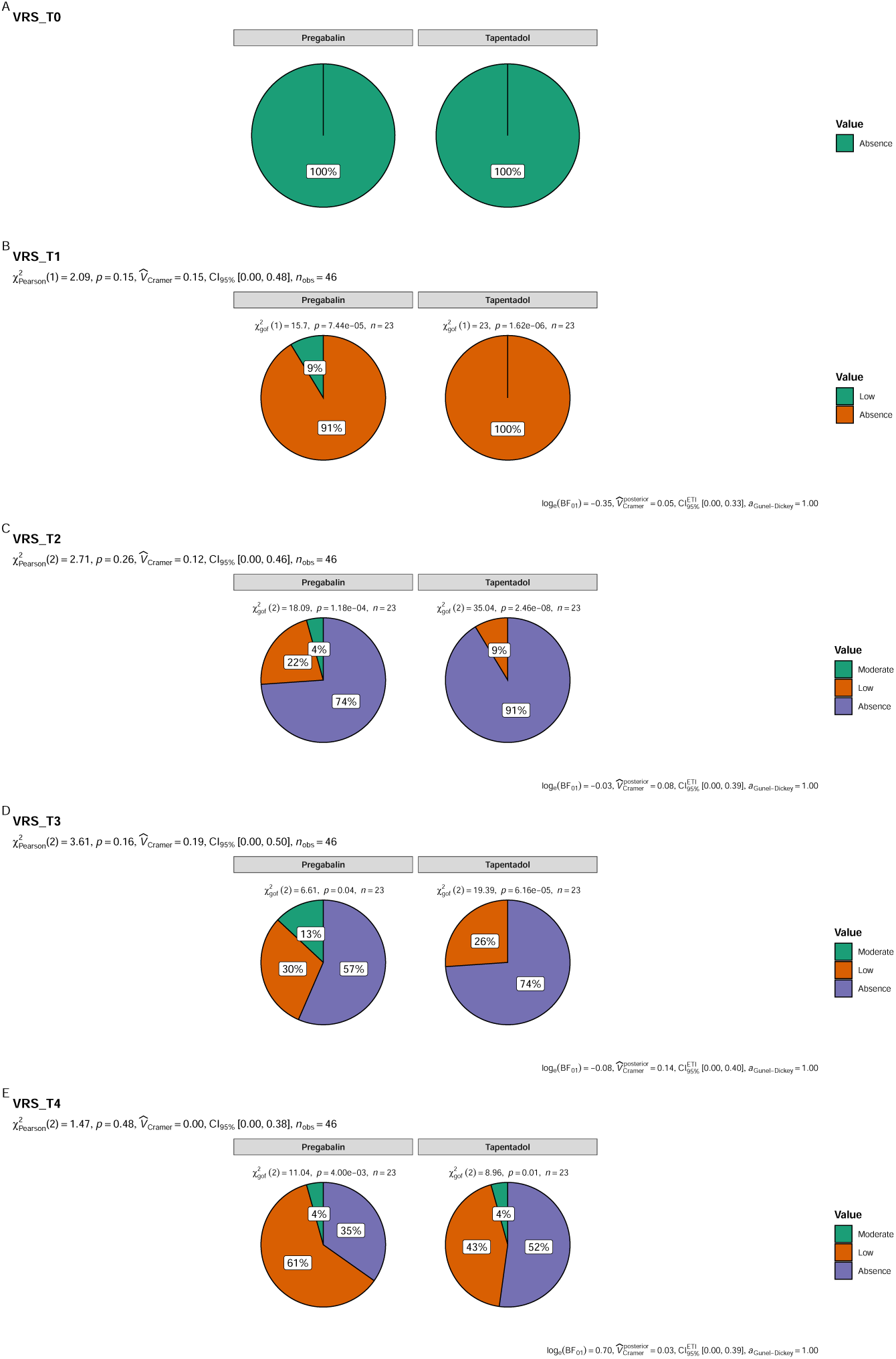
Verbal Rating Scale (VRS) pain categories over time by treatment group. Stacked bar charts show the proportion of patients reporting absence, low, or moderate pain at each timepoint. All patients reported absence of pain at T0. The proportion reporting any pain was consistently higher in the pregabalin group from T1 onward, with 13.0 % reporting moderate pain at T3 compared with 0 % in the tapentadol group. TAP = tapentadol; PREG = pregabalin; VRS = Verbal Rating Scale.

### 4.5. Secondary Outcome: Rescue Analgesia

Rescue analgesia was required by 1 patient (4.3%) in the tapentadol group and 5 patients (21.7%) in the pregabalin group during the two-hour PACU observation period. All rescue doses were administered at T2, T3, or T4, coinciding with the timepoints of maximal between-group divergence in NRS scores. Total morphine consumption reflected the same pattern, with the pregabalin group requiring approximately five times the cumulative rescue opioid dose of the tapentadol group, though between-group differences in total morphine consumption did not reach statistical significance given the small number of rescue events.

**Figure 4:**
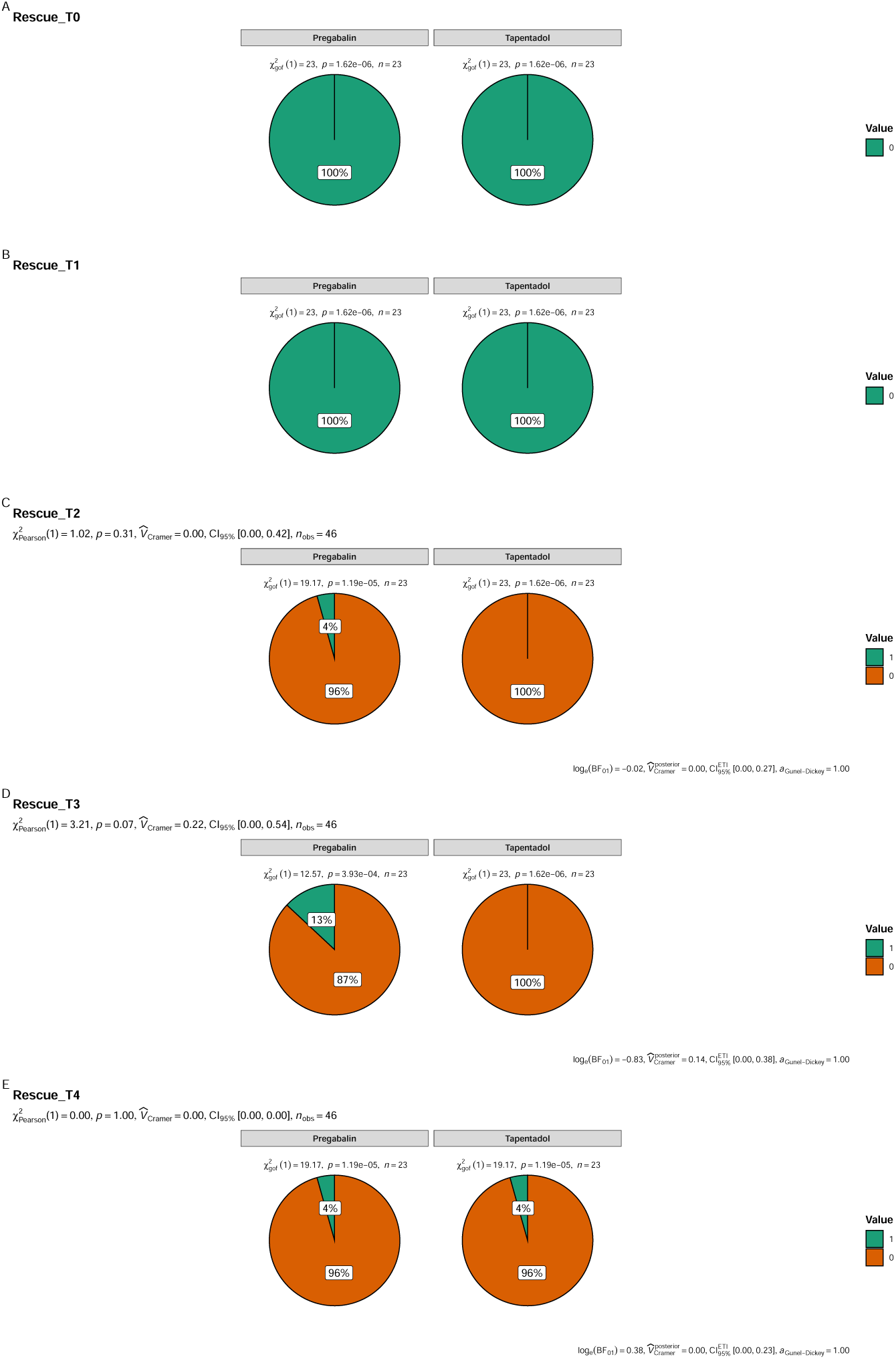
Rescue analgesia requirements by treatment group. Proportion of patients requiring morphine rescue (4 mg IV) during the two-hour PACU observation period. Rescue analgesia was required by 4.3 % of tapentadol-premedicated patients (1/23) compared with 21.7 % of pregabalin-premedicated patients (5/23). TAP = tapentadol; PREG = pregabalin; PACU = post-anesthesia care unit.

### 4.6. Safety

The incidence of nausea and vomiting was identical in both groups (17.4% in each group), with no statistically significant difference (*p* = 1.0). No anaphylactic or hypersensitivity reactions were observed in either group. No patients required advanced airway management, vasopressor support, or unplanned intensive care admission.

## 5. Discussion

This randomized controlled trial evaluated the efficacy of oral premedication with tapentadol compared to pregabalin for acute postoperative pain in patients undergoing elective lower limb surgery under neuraxial anesthesia. The study demonstrated that tapentadol was associated with significantly lower pain scores and reduced rescue analgesia requirements compared with pregabalin. By 90 minutes post-arrival, the observed difference in NRS was –0.91 (p = 0.006, Cohen’s d = –0.96). Rescue morphine was required in 4.3% of tapentadol-treated patients versus 21.7% of pregabalin-treated patients. Both agents were well tolerated, with identical rates of nausea/vomiting (17.4%) and no hypersensitivity reactions.

### 5.1. Summary of Key Findings

Tapentadol premedication provided superior analgesia compared to pregabalin at the 90-minute timepoint, with a large effect size (Cohen’s d = 0.96). The proportion of patients requiring rescue analgesia was five times lower in the tapentadol group. The temporal pattern of pain divergence — absent at T0 and T1, emerging at T2, and most pronounced at T3 — is consistent with the pharmacokinetic profile of immediate-release tapentadol, which reaches peak plasma concentration within 3–6 hours of oral administration and maintains stable analgesic effect from the third dose onward [9, 16]. The 72-hour premedication regimen was designed to achieve steady-state concentrations, and the analgesic trajectory supports this rationale. Tapentadol’s dual mechanism (*µ*-opioid agonism and norepinephrine reuptake inhibition) may offer advantages over pregabalin’s pure presynaptic calcium-channel modulation in the context of orthopaedic surgery, where sustained nociceptive input from periosteum is substantial [5, 8].

The overall Group *×* Time interaction in the primary LMM did not reach conventional statistical significance (p = 0.142), which is consistent with the limited power of the sample size. The borderline-significant interaction in the continuous-time sensitivity model (p = 0.053) and the large-effect post-hoc contrast at T3 (d = 0.96) together suggest that the lack of a significant overall interaction reflects insufficient power rather than absence of a true effect. These findings provide effect size and variance estimates that can inform future confirmatory trials.

### 5.2. Comparison with Previous Studies

The present findings extend the existing literature on preemptive analgesia with gabapentinoids and atypical opioids. Pregabalin’s perioperative analgesic efficacy has been demonstrated in laparoscopic cholecystectomy and anterior cruciate ligament repair [6, 7], but comparative data against tapentadol in the orthopaedic lower limb context are absent. Our study provides the first direct comparison of these two agents as oral premedication in lower limb surgery.

The comparable adverse event profiles observed here — specifically, identical rates of nausea and vomiting (17.4% in each group) — are consistent with the known tolerability advantage of tapentadol over traditional opioids and are reassuring in the context of a preoperative oral regimen [9, 17]. The absence of hypersensitivity reactions in either arm supports the safety of both agents at the doses employed.

### 5.3. Strengths and Limitations

This study has several strengths. The double-blind, randomised design with a formal sample size calculation provides a rigorous comparison of the two agents. The use of neuraxial anaesthesia standardised the surgical and anaesthetic conditions. Protocol adherence and retention were 100%, supporting the feasibility of this intervention in clinical practice.

Several limitations must be acknowledged. First, the sample size (n = 23 per group) limits statistical power for detecting interaction effects; therefore, the results should be interpreted cautiously. Second, pain assessment was limited to the first 120 minutes of PACU recovery; longer follow-up would be needed to assess the full analgesic trajectory and the risk of chronic pain. Third, the heterogeneity of orthopaedic procedures introduces variability in nociceptive stimulus. Fourth, the trial was conducted at a single centre, limiting generalisability. Fifth, https://ClinicalTrials.gov registration was completed after data collection because access to an international registry was not available at the time of study initiation; this is disclosed transparently and does not affect the integrity of the data.

### 5.4. Implications for Future Research

The observed variance (SD = 1.2) provides a concrete basis for sample size estimation for future trials. Based on this variance, a non-inferiority trial with a margin of 1 NRS point, *α* = 0.05 (two-sided), and power = 80% would require approximately 40 patients per group (80 total). Future trials should consider longer follow-up, stratification by surgical procedure, and inclusion of patient-centred outcomes such as quality of recovery or functional status.

## 6. Conclusions

In this randomized controlled trial of 46 patients undergoing lower limb surgery under neuraxial anesthesia, 72-hour oral premedication with tapentadol 100 mg/day provided superior postoperative analgesia compared with pregabalin 75 mg/day at the clinically critical 90-minute PACU timepoint, with a large effect size (Cohen’s d = 0.96) and a fivefold reduction in rescue analgesia requirements (4.3% vs. 21.7%). Both agents were well tolerated, with identical rates of nausea and vomiting and no hypersensitivity events in either arm. These findings support the use of tapentadol as a premedication strategy in lower limb orthopaedic surgery and provide a basis for a larger confirmatory trial.

## 7. Authors’ Contributions

ME: conceptualization, data curation, formal analysis, investigation, methodology, project administration, resources, supervision, validation, writing-reviewing and editing; PL: data curation, formal analysis, methodology, project administration, software, supervision, validation, visualization, writing-reviewing and editing; MD: conceptualization, funding acquisition, investigation, methodology, project administration, resources, supervision, validation, writing-reviewing and editing.

All authors have read and approved the final manuscript.

## 8. Acknowledgments

The authors acknowledge the *Instituto de Seguridad y Servicios Sociales de los Trabajadores del Estado Hospital Regional “General Ignacio Zaragoza”*, its Department of Anesthesiology, Dr. Miguel Pineda Sánchez, M.D.; Dr. Humberto Pineda Domínguez, M.D.; Dr. Dalia Vásquez Vásquez, M.D.; and the *Universidad Nacional Autónoma de México*.

## 9. Declaration of Interest

The authors declare that they have no conflict of interest.

## 10. Funding

Funding and equipment were provided by the *Instituto de Seguridad y Servicios Sociales de los Trabajadores del Estado Hospital Regional “General Ignacio Zaragoza”*.

## 11. Data Availability

Deidentified individual participant data will be made available following article publication, on GitHub <https://github.com/phabel-LD>, along with all analysis code.

## Notes

### Competing Interest Statement

The authors have declared no competing interest.

### Clinical Trial

NCT07587645

### Author Declarations

The Ethics Committee of the Hospital Regional "General Ignacio Zaragoza", Instituto de Seguridad y Servicios Sociales de los Trabajadores del Estado gave ethical approval for this work.

### Summary of Updates

This revised version has been updated to reflect the original study design as a randomized controlled trial. The term "pilot" has been removed from the title, abstract, and manuscript. The sample size calculation (two-proportion superiority, 23 patients per group, 46 total) has been added to the Methods section. Feasibility and exploratory language have been removed. The running head has been corrected to "Escalona-Arroyo et al., Tapentadol vs Pregabalin: RCT."

